# Vaccine-preventable diseases other than tuberculosis, and homelessness: A systematic review of the published literature, 1980 to 2020

**DOI:** 10.1101/2020.10.28.20220335

**Authors:** Tran Duc Anh Ly, Sergei Castaneda, Van Thuan Hoang, Thi Loi Dao, Philippe Gautret

**Affiliations:** Aix Marseille Univ, IRD, AP-HM, SSA, VITROME, Marseille, France; IHU-Méditerranée Infection, Marseille, France; Family Medicine Department, Thai Binh University of Medicine and Pharmacy, Vietnam; Pneumology Department, Thai Binh University of Medicine and Pharmacy, Vietnam

**Author notes:** **Corresponding author:** Philippe Gautret, VITROME, Institut Hospitalo-Universitaire Méditerranée Infection, 19-21 Boulevard Jean Moulin 13385 Marseille Cedex 05, France.

**Keywords:** Homelessness, outbreak, vaccine-preventable diseases, immunization, vaccination recommendation

## Abstract

**Background:** Homelessness may result in the breakdown of regular health services, including routine vaccination programmes. A literature review was conducted to describe vaccine-preventable diseases (VPD) other than tuberculosis in homeless populations and to summarize vaccination recommendations in homeless people.

**Methods:** We followed Preferred Reporting Items for Systematic reviews and Meta-Analyses (PRISMA) guidelines. We searched peer-reviewed literature published in English, French or Spanish reporting the outbreak of VPD or VPD prevalence in both infant and adult homeless populations published between 1980 and 2020, using PubMed/Medline, SciELO, Google Scholar, and Web of Science databases. Relevant information from the studies was charted in Microsoft Excel and results were summarised using a descriptive analytical method.

**Results:** Seventy-five articles were included. A high prevalence of past HBV and HAV infections were observed through serosurveys, mostly in high income countries or high-middle income countries (USA, Canada, France, Iran or Brazil). Nine outbreaks of HAV infection were also reported, with lethality rates ranging from 0-4.8%. The studies discussed numerous risk factors positively associated with HBV infection, including older age, homosexual or bisexual practice, injected drug use (IDU), and, with HAV infection including IDU, having sexual partner(s) with a history of unspecified hepatitis, insertive anal penetration, or originating from a country with a high prevalence of anti-HAV antibody. Eleven outbreaks of pneumococcal infection affecting homeless persons were reported in Canada and USA, with lethality rates from 0-15.6%. Five diphtheria outbreaks were reported. Vaccination status was rarely documented in these studies.

**Conclusions:** The literature suggests that homeless populations generally experience a higher VPD burden and lower immunisation rates. The findings suggest the need for a national vaccination programme and planning for delivering vaccines in this population.

## 1. Introduction

In addition to mental illness and unintentional injuries, homeless people are exposed to communicable diseases, which may lead to outbreaks that can become serious public health concerns [1]. High rates of infectious disease have been recorded in the literature among homeless people, including respiratory tract infections [2], gastrointestinal infections [3], skin infections, arthropod-borne diseases [4], and blood-borne and sexually transmitted infections [1].

The prevalence of these infectious diseases among the homeless varies greatly, depending on exposure factors. Generally, frequent alcohol and tobacco consumption or illicit drug use significantly impair the health status of homeless individuals [1]. Those sleeping on the street, outdoors in vehicles or in abandoned buildings are at high risk of food-borne disease (as the result of exposure to unhygienic environments) [3], human louse-transmitted disease [5], and of blood-borne and sexually transmitted infections (as the result of promiscuity and lack of hygiene, injected drug use and unprotected sexual practices) [6-7]. In addition, living in overcrowded accommodations, poor environmental conditions (poor ventilation, lack of sufficient food) and lack of continuity of care (omission of preventive measures, delayed diagnosis, interrupted or poor quality treatment) may increase vulnerability to infection, including transmissible disease. Consequently, when outbreaks occur in these settings, they spread rapidly, often with a high probability of prolonged transmission [1]. Evidence suggests that vaccination is still the most effective strategy for preventing infectious disease, resulting in significant reductions in illness, disability and death from several VPD [8]. Implementing routine vaccination may not only ensure long-term protection against VPD through the progressive increase of population immunity, limiting the number of preventable deaths among homeless people during an outbreak [3], but also protects vulnerable members of the general population who have not received all vaccines, such as the immunocompromised and the very young [9].

TB in homeless people has been extensively described in several recent review papers [10-13]. This scoping review of the published literature aims to describe an outbreak of VPD or VPD prevalence other than TB in both an infant and adult homeless population during the period 1980-2020. In addition, vaccination recommendations in homeless people were reviewed.

## 2. Materials and Methods

### 2.2 Search strategy

The systematic review followed the Preferred Reporting Items for Systematic Reviews and Meta-Analyses (PRISMA) guidelines (http://www.prisma-statement.org). Using Harzing’s Publish or Perish software (version 7.0), we searched the published literature from 1980 to 04 September 2020 in PubMed/Medline (in English), Scopus (in English), Web of Science databases (in English), SciELO (in English and Spanish), and Google Scholar (in French). Search strings were tailored to each database. Article references were scanned for additional articles. References were managed by Endnote software (version 9.3), and relevant information from the studies was charted in Microsoft Excel. Data table elements were created and were reviewed by all authors to ensure consistency of information extraction. Combinations of the following English search terms were used:

> #1: “homeless” OR “homelessness” OR “street people”
>
> #2: “diphtheria” OR “*Corynebacterium diphtheriae*” OR “tetanus” OR “*Condyloma accuminata”* OR “polio” OR “poliovirus” OR “poliomyelitis” OR “pertussis” OR “whooping cough” OR “*Bordetella pertussis*” OR “*Haemophilus influenzae* type b” OR “hepatitis B” OR “pneumococcal” OR “*Streptococcus pneumoniae*” OR “meningococcal” OR “meningococcus” OR “*Neisseria meningitidis*” OR “mumps” OR “measles” OR “rubella” OR “Human papillomavirus” OR “influenza” OR “flu” OR “hepatitis A” OR “shingles” OR “Varicella” OR “chickenpox” OR “typhoid” OR “Salmonella” OR “Rotavirus” OR “cholera” OR “*Vibrio cholerae* “OR “yellow fever” OR “rabies” OR “lyssaviruses” OR “Japanese encephalitis” OR “leptospirosis” OR “Leptospira”
>
> #3: #1 AND #2

Combinations of the search terms in Spanish and French are shown in Supplementary data.

### 2.3 Inclusion and exclusion criteria

For inclusion, an article had to fulfil the following criteria: articles (1) written in English (focused language), French, Spanish, or Portuguese languages (if any); (2) published in peer-reviewed journals or unpublished data in our laboratory regarding the homeless population in Marseille, France; (3) reporting outbreaks of VPD or VPD prevalence in an infant or adult homeless population. Articles were excluded if the studies were reviews, case reports, or reported only vaccination rates. An additional manual search was conducted by reviewing reference lists of papers retrieved through the electronic search.

## 3. Results

As presented in Figure 1, the search identified a total of 9499 records, with an additional 2 records being unpublished data in our laboratory, resulting in 75 articles/reports that fulfilled the eligibility criteria (Figure 1). Seventy-one were written in English, one in French and two in Spanish. One article written in Portuguese was found when searching abstracts in English and was also included. The main findings of these articles are presented in Table 1.

**Table 1.**
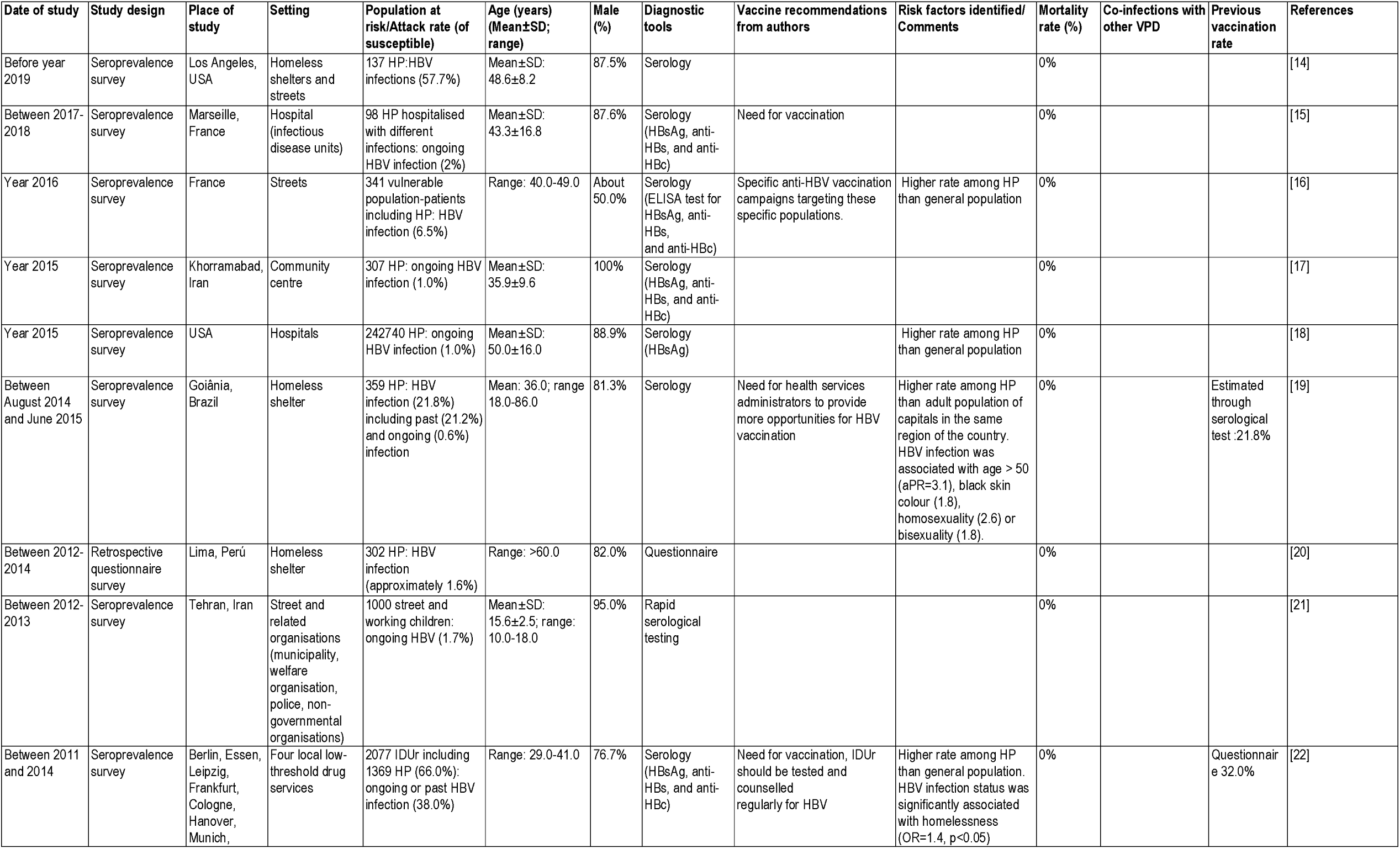
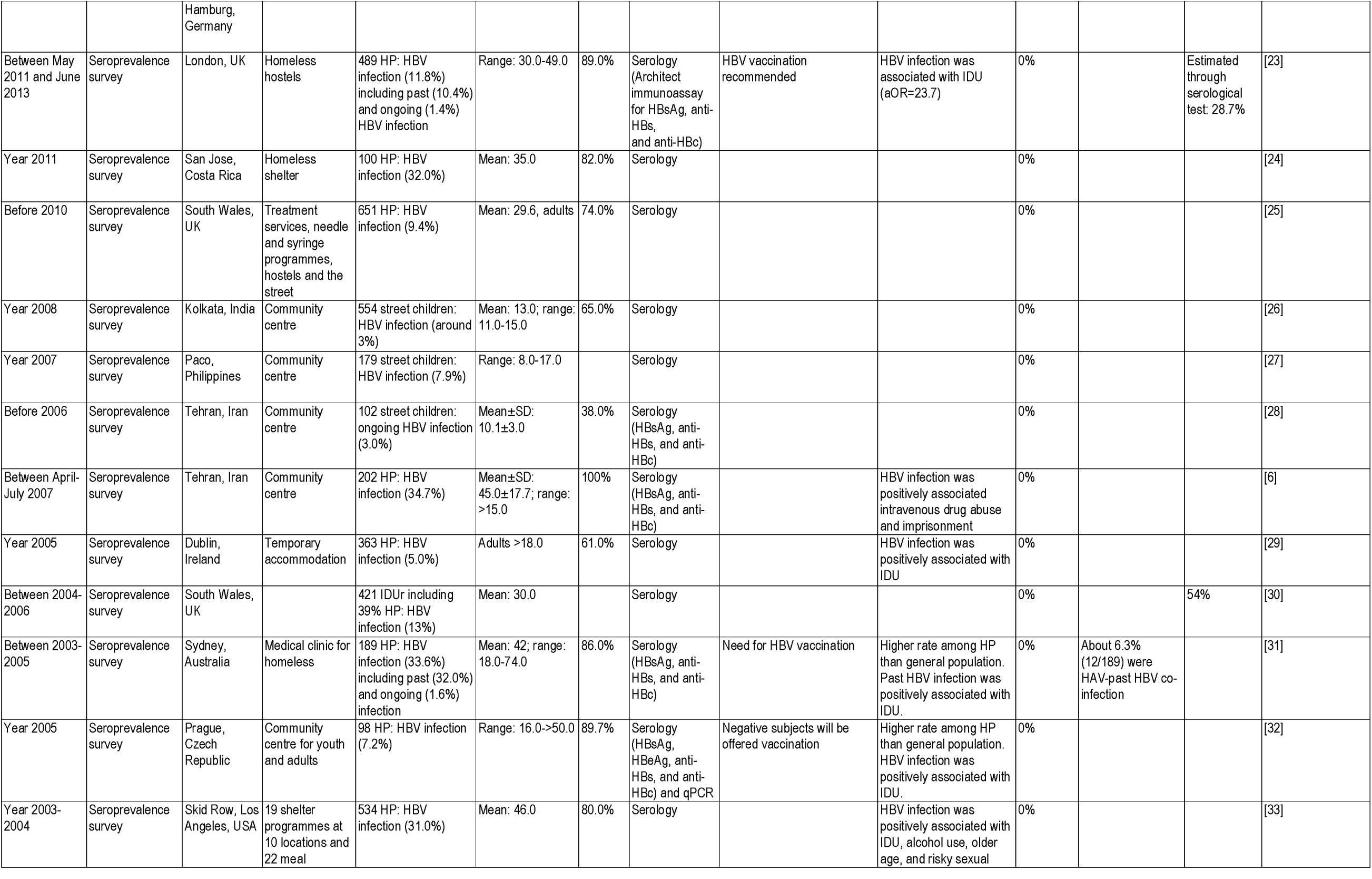
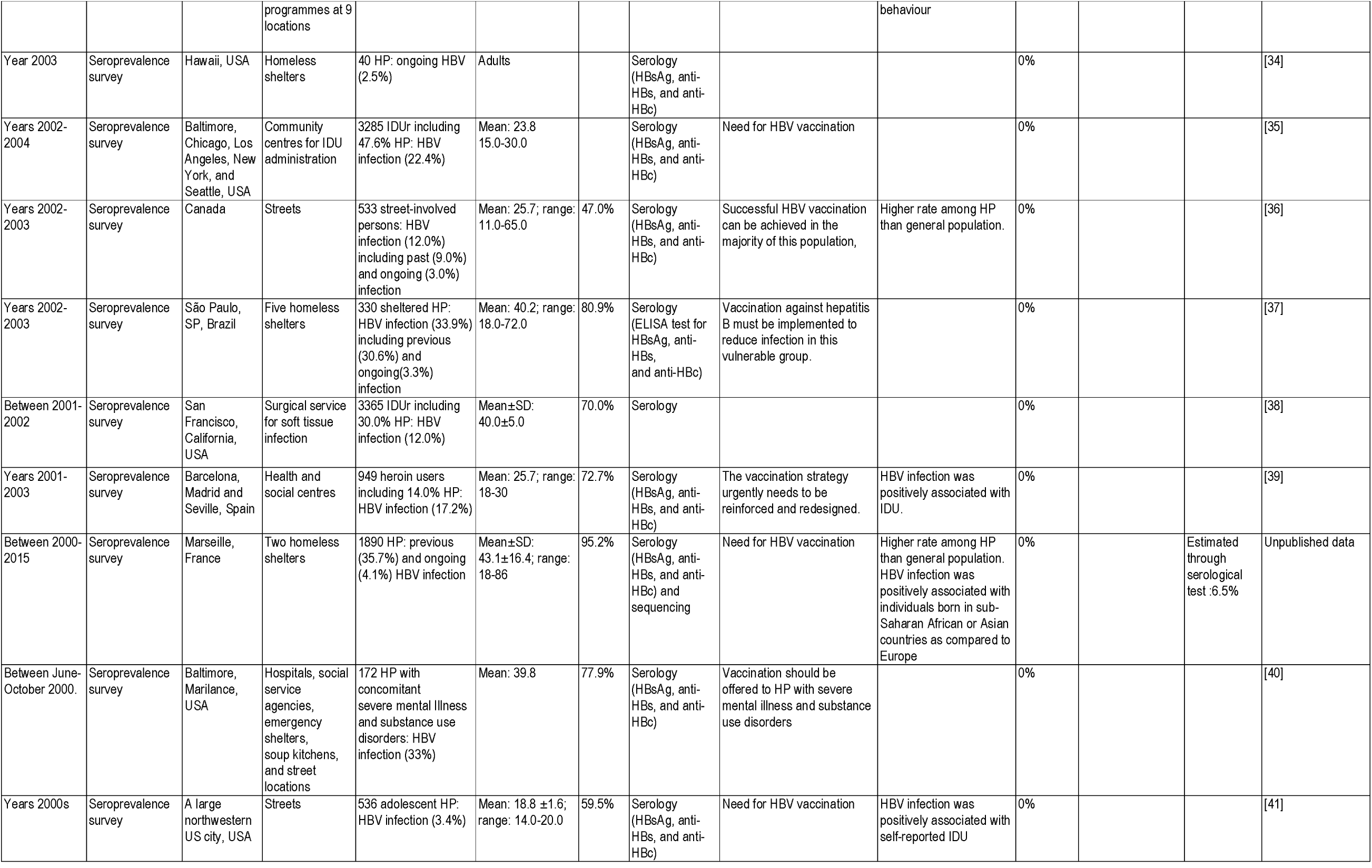
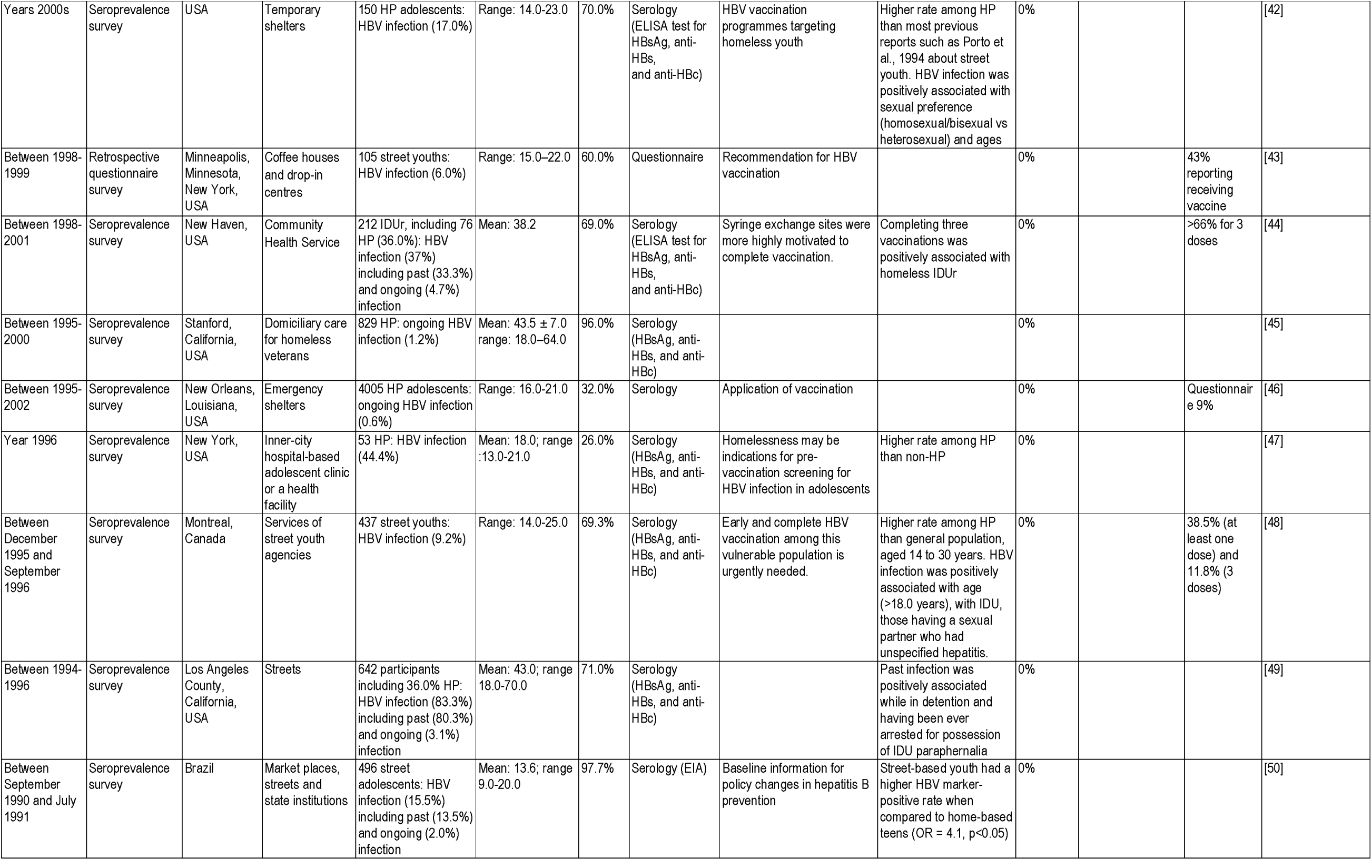
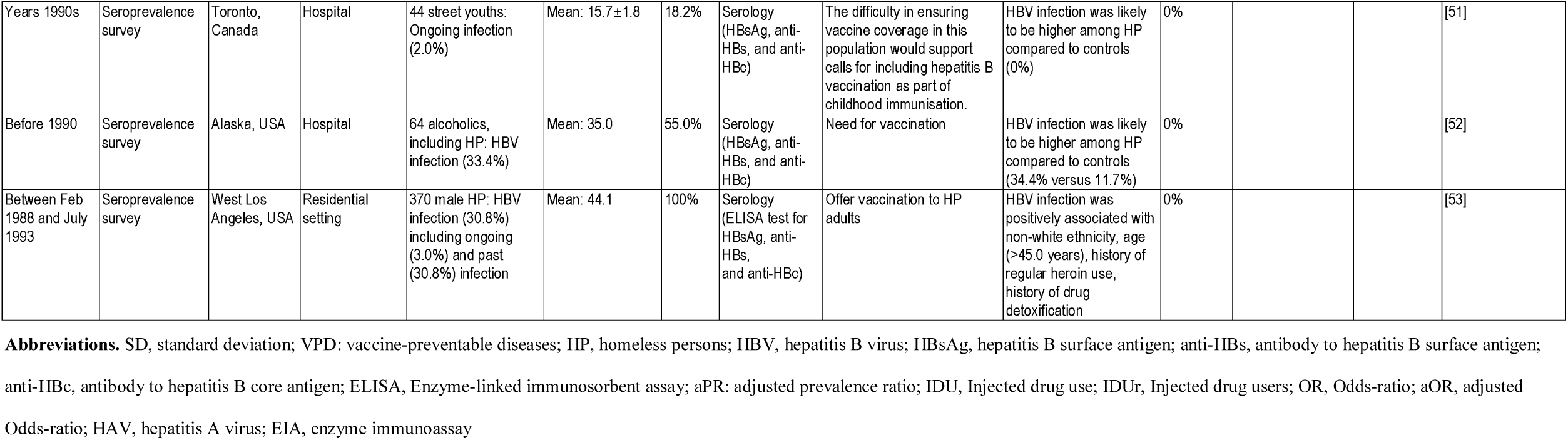
Hepatitis B infection among homeless persons identified through record review studies.

**Figure 1.**
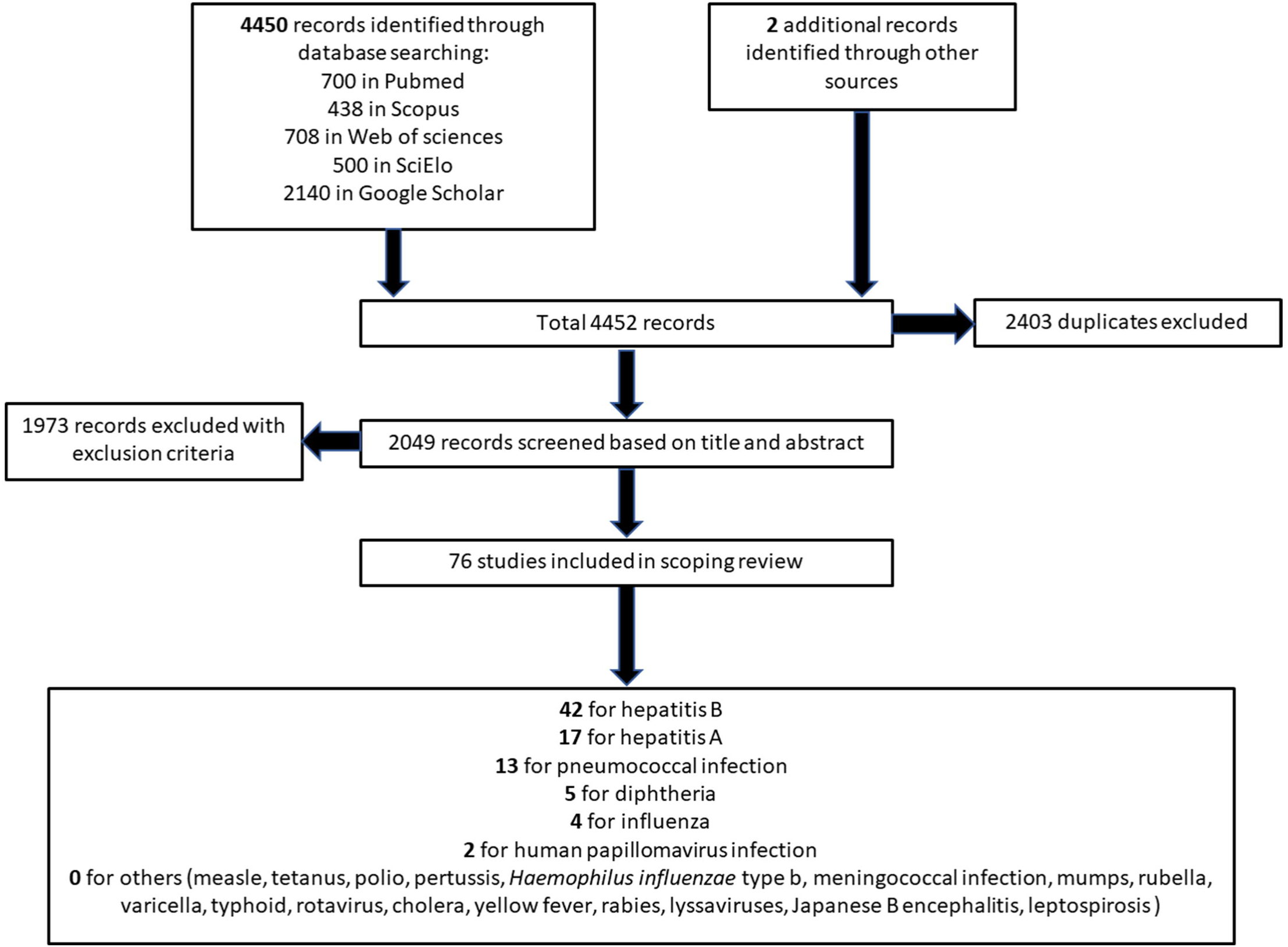
Flow diagram of included and excluded records. * Two unpublished data sets in our laboratory.

### Hepatitis B (Table 1)

Forty-two hepatitis B articles reporting on vulnerable populations, notably including homeless people, were retrieved (including an unpublished data set) [6, 14-53]. Overall, 269,750 individuals were enrolled between 1988 and 2019. Thirty studies were conducted in adults (mean age: 25-49 years, range 16-86 years) [6, 14, 15, 17-25, 29-34, 37-40, 44-47, 49, 51-53], five studies were conducted in street youth (mean age:10-16 years, range 8-18 years) [16, 26-28, 51], and seven in both children and adults (mean age: 14-26 years, range 9-65 years) [35, 36, 41-43, 48, 50]. Studies were conducted in the USA (n=17) [14, 18, 33-35, 38, 40-47, 49, 52, 53], Iran (n=4) [6, 17, 21, 28], Canada [36, 48, 51], Brazil [19, 37, 50] France [15, 16] (including unpublished data), the UK [23, 25, 33], (n=3 per country), Australia [31], Costa Rica [24], Czech Republic [32], Germany [22], India [26], Ireland [29], Peru [20], Philippines [27] and Spain [39] (n=1 per country). Forty studies were seroprevalence surveys [6, 14-19, 21-42, 44-53]. The prevalence of anti-HBc antibodies ranged from 10.4% to 80.3% (past infection) and that of HBsAg ranged from 0.6% to 4.7% (ongoing infection). Two retrospective questionnaire surveys reported rates of 1.6% and 6% for HBV infection among participants [20, 43]. HBV infection among the homeless population was significantly higher than in non-homeless individuals included in the studies, or than in the general population, as assessed in studies conducted at the same period (n=14, including one unpublished data set) [16, 18, 19, 22, 31, 32, 36, 42, 47, 48, 51-53].

HBV infection among the homeless population was shown to be positively associated with older age [19, 42, 48], with homosexual or bisexual practices, having a sexual partner(s) with a history of unspecified hepatitis or insertive anal penetration [19, 42, 48], having black skin colour [19], reporting injected drug use (IDU) [6, 22-24, 29-33, 35, 38, 39, 41, 44, 48, 49], and alcohol use [33]. One study conducted among 189 homeless individuals in Australia reported that 6.3% of individuals with a past HBV infection also had a past hepatitis A (HAV) infection [31]. No hepatitis B–related deaths were reported. Five studies reported vaccination status against HBV infection through a questionnaire, with rates of 9%-54% (at least one dose) and 11.8%->66% (3 doses) [22, 30, 43, 44, 46, 48], and three studies showed immunisation rates of 21.8%-38.7% through serological tests [19, 23]. Anti-HBV vaccination was strongly recommended by all authors for high risk groups such as street youth or injected drug users (n=24 studies, including unpublished data) [15, 16, 19, 22, 23, 31, 32, 35-37, 39-44, 46-48, 50-53].

### Hepatitis A (Table 2)

Seventeen hepatitis A articles conducted among homeless populations fulfilled the eligibility criteria [31, 35, 36, 54-67]. Overall, 10,552 individuals were enrolled between 1991 and 2018. Nine studies were conducted in adults (mean age: 19-45 years, range 16-74 years) [31, 55-57, 59, 61, 62, 64, 65], and eight in populations comprising children and adults (mean age: 25-42 years, range 0-90 years) [35, 36, 54, 58, 60, 63, 66, 67]. Studies were conducted in the USA (n=7) [35, 54, 56-68, 61, 64], Canada (n=4) [36, 55, 65, 66], Brazil [67], the UK [63], Czech Republic [60], The Netherlands [62], Iran [59] and Australia (n=1 per country) [31].

**Table 2.**
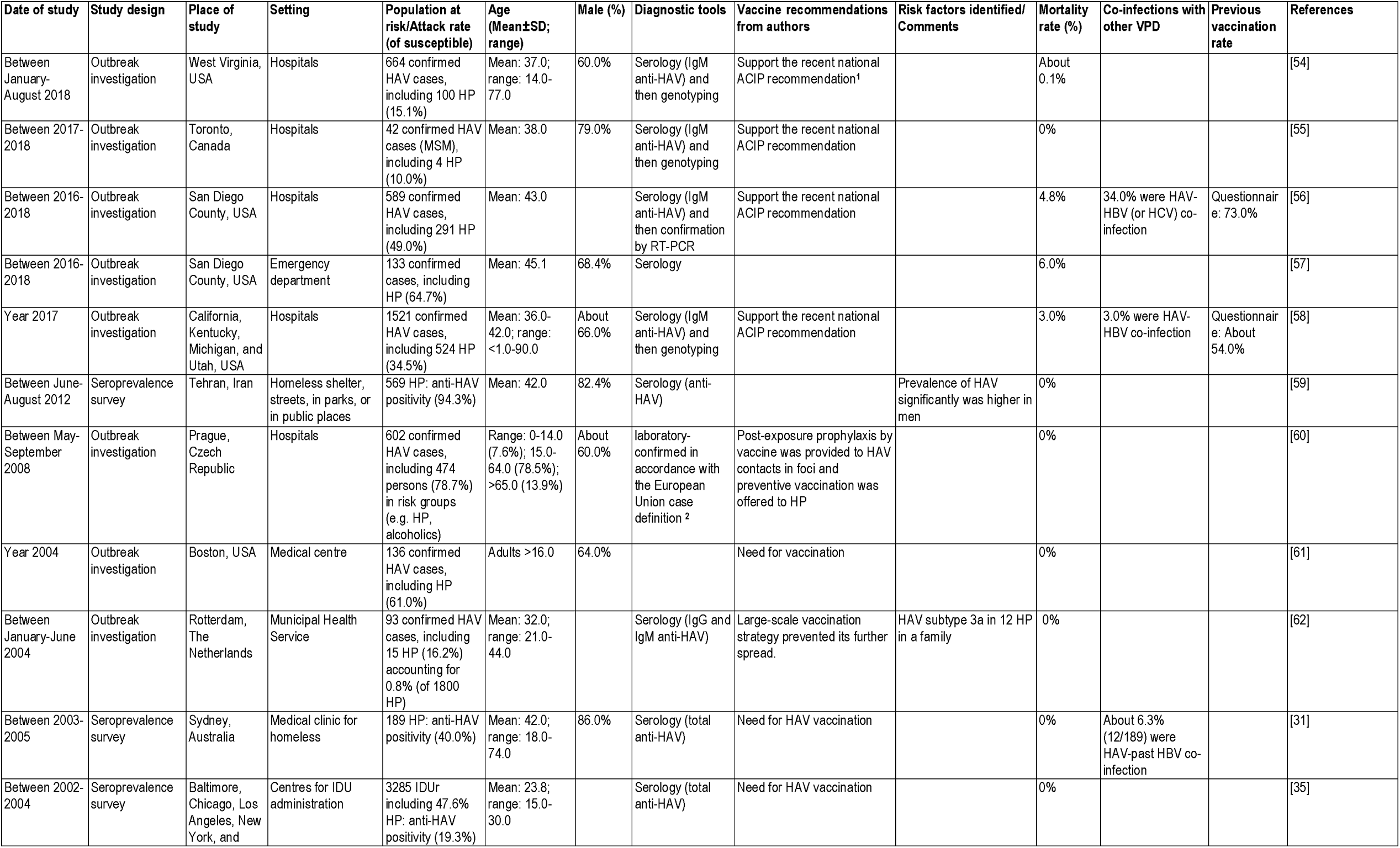
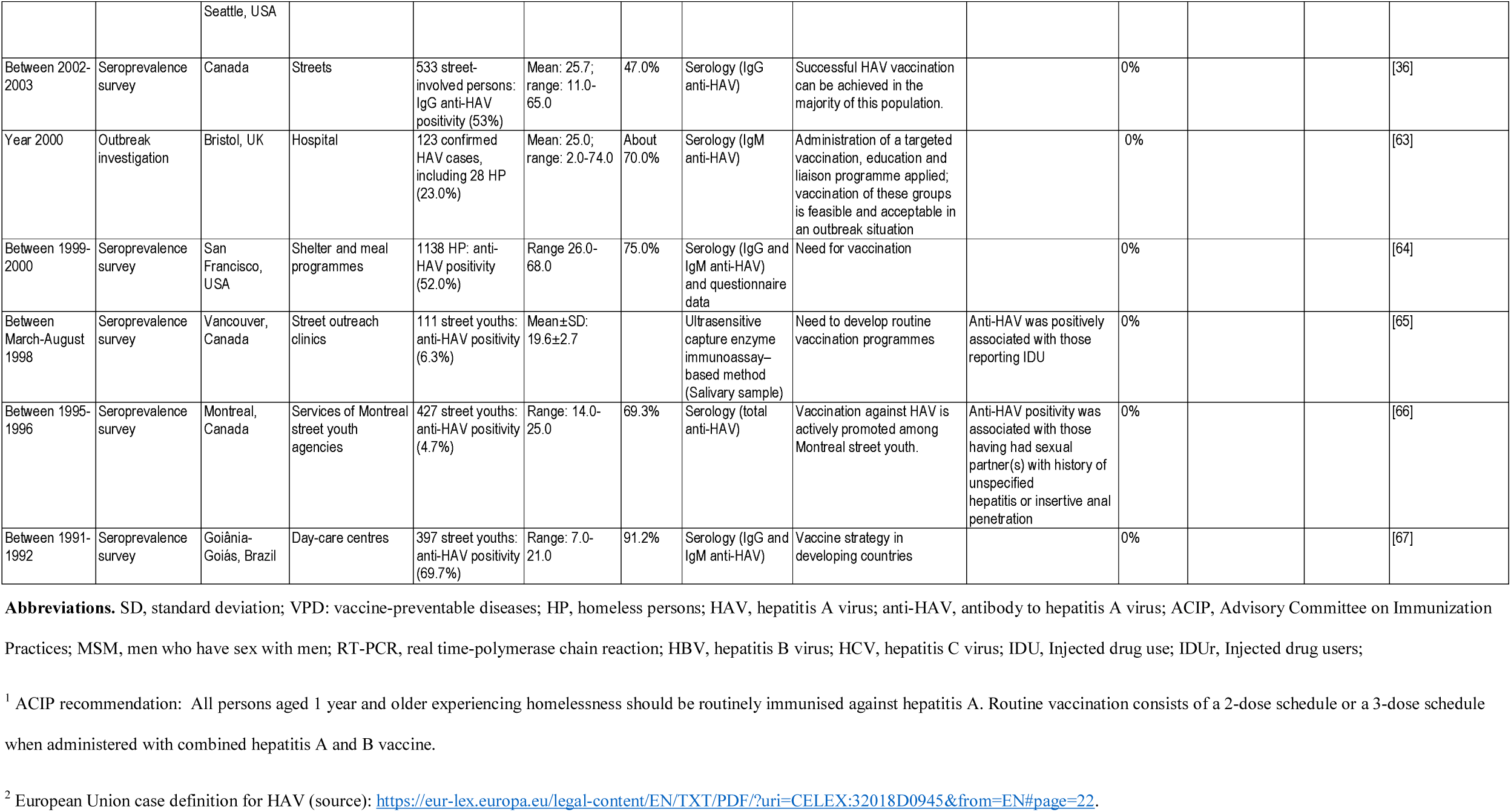
Hepatitis A infection among homeless persons identified through record review studies.

Eight surveys addressed the seroprevalence of total anti-HAV antibodies among 6649 individuals: six revealed a high prevalence of 19.3%-69.7% among homeless persons in Brazil, Canada and the USA [31, 35, 36, 59, 64, 67], while two studies showed lower seroprevalence rates of 4.7%-6.3% among Canadian street youth, but with a high prevalence of risk factors for infection [65, 66]. Three studies reported risk factors for a high prevalence of HAV infection (reporting IDU, having sexual partner(s) with history of unspecified hepatitis, male subjects reporting insertive anal penetration or those originating from a country with a high anti-HAV prevalence) [35, 65, 66]. Other studies described nine outbreaks of Hepatitis A in several cities in the UK, The Netherlands, Czech republic and the USA; the numbers of confirmed cases in hospitals ranged from 42 to 1521 per outbreak (resulting in a total of 3903 outbreak-associated cases), with a high prevalence of 15.1%-78.0% being homeless persons [54-58, 60-63]. HAV-associated deaths ranged from 0-4.8% during outbreaks. Rates of co-infection with HAV and HBV (or HCV) were 3.0%, 6.3% and 34.0% in three studies [31, 55, 58]. No data about previous vaccination rates were available. Vaccination of homeless populations against HAV was strongly recommended in 15 studies [31, 35, 36, 54-56, 58, 60-67].

### Pneumococcal infections (Table 3)

Thirteen studies concerning pneumococcal infections in homeless people were retrieved between 1988 and 2017 (including one unpublished data set) [2, 68-78]: 12 studies were conducted in adults (mean age: 19-54 years, range 16-78 years) and one in a population comprising children and adults (mean age: 42 years, range 0-90 years). Most studies were conducted in Canada (n=8) [68, 70-76], followed by France (n=3, including one unpublished data set) [2, 78], and the USA (n=2) [69, 77]. Two molecular prevalence surveys reported rates of 12.4% and 15.5% for pneumococcal nasopharyngeal carriage among a total of 575 sheltered homeless persons in Marseille, France between 2015-2018 (including one unpublished data set) [2], and prevalence was positively associated with respiratory symptoms and signs. Other studies described eleven outbreaks of invasive pneumococcal disease (IPD) occurring in different cities in Canada, the USA and France; of those, two studies reported 153 homeless individuals hospitalised with IPD, which accounted for 1.4%-8.4% of all homeless people present in two cities, Toronto (Canada) and Anchorage (USA) [76, 77]; nine other studies reported a high proportion (4.7%-48.8%) of homelessness among 4742 individuals hospitalised with IPD [68-75, 78]. Homeless individuals with IDP were typically younger [73, 77], more often male [73], smokers[73, 75, 76], alcohol abusers [73, 75, 77], illegal drug users [73, 75], and had a primary diagnosis of pneumonia [73], HIV infection or liver disease [76] when compared with non-homeless individuals. Of nine studies, two characterising the serotype of isolated *Streptococcus pneumoniae* strains showed that serotypes 1, 4, 5, 8 and 12F were positively associated with homelessness in Canada [68, 70, 72]. A serotype 1 IPD outbreak was reported among sheltered homeless individuals in Paris, France between 1988-1989 [78]. Serotype 4 outbreaks were also positively associated with homelessness in New Mexico, USA [69]. The mortality rate was 0%-15.6% among infected homeless people during outbreaks. By using a questionnaire, previous vaccination rates among the homeless population were reported to be 3.1%-37.0% (including one unpublished data set) [68, 70, 76, 78]. The pneumococcal vaccine was recommended for homeless populations in all articles.

**Table 3.** Other vaccine-preventable diseases among homeless persons identified through record review studies.

| Date of study | Study design | Place of study | Setting | Population at risk/Attack rate (of susceptible) | Age (Mean±SD; range) | Male (%) | Diagnostic tools | Vaccine recommendations from authors | Risk factors identified/ Comments | Mortality rate (%) | Co-infections with other VPD | Previous vaccination rate | References |
| --- | --- | --- | --- | --- | --- | --- | --- | --- | --- | --- | --- | --- | --- |
| **A. Pneumococcal infection** |  |  |  |  |  |  |  |  |  |  |  |  |  |
| Year 2018 | Molecular prevalence survey | Marseille, France | Two homeless shelters | 98 HP: Pneumococcal nasopharyngeal carriage (15.5%) | Mean±SD: 39.3 ± 17.6 | 100% | qPCR | Need to increase vaccination rate | Pneumococcal carriage prevalence was likely to be higher than control groups | 0% |  | Questionnaire: 3.1% | Unpublished data |
| Autumn and winter of 2016–2017 | Outbreak investigation | Victoria, British Columbia, Canada | Hospitals | 84 HP with confirmed IPD | Mean: 56.3 | 61.0% | Culture isolation and serotyping (Gram stain, colony morphology on blood agar, bile solubility, susceptibility to optochin) | Pneumococcal vaccination coverage is needed among this marginalised population. | Positive association between serotype 4 IPD and homelessness (OR=4.9; p<0.01) | 11.9% |  | Questionnaire: 13.0% | [68] |
| Between 2015–2017 | Molecular prevalence survey | Marseille, France | Two homeless shelters | 477 HP: Pneumococcal nasopharyngeal carriage (12.4%) | Mean±SD: 43.6±16.0; range: 18.0–84.0 | 100% | qPCR | Need to increase vaccination rate | Pneumococcal carriage was positively associated with respiratory symptom presentation. | 0% | 0.2% were *S. pneumococcus*-influenza virus co-carriage |  | [2] |
| Between 2010–2018 | Outbreak investigation | California, Colorado, and New Mexico, USA | Hospitals | 325 serotype 4 cases among adults, including HP (36.0%) | Adults ≥ 17.0 |  | Culture isolation and whole-genome sequencing | Future vaccine policy discussions should include homelessness as an indication for pneumococcal vaccines. |  |  |  |  | [69] |
| Between September 2009 and January 2011 | Outbreak investigation | Winnipeg, Canada | Hospitals | 169 patients with confirmed IPD, including 20 HP (11.8%) | Mean: 42.0; range: 0–94.0 | About 58.0% | Culture isolation, PFGE, MLST, MLVA | Need to vaccinate unvaccinated individuals who are homeless or use illicit drugs | Serotype 12F patients were more likely to report homelessness (OR=12.7; p<0.05). | 0% |  | Questionnaire: 37.0% | [70] |
| Between 2005–2009 | Outbreak investigation | Alberta, Canada | Hospitals | 1112 patients with confirmed IPD, including 85 HP (7.6%) | Mean±SD: 45.4±22.5 | About 59.3% | Culture isolation from site such as blood, CSF, pleural fluid, biopsy tissue, joint aspiration, pericardial fluid, or peritoneal fluid | Need for vaccination programme for HP |  | About 11.3% |  |  | [71] |
| Between 2005–2007 | Outbreak investigation | Calgary, Canada | Hospitals | 207 patients with confirmed IPD (strain ST5 or ST8), including HP (48.8%) | Adults ≥ 16.0 |  | Gram stain, colony morphology on blood agar, bile solubility, susceptibility to optochin and pneumococcal antibody agglutination and susceptibility testing (broth microdilution). | Vaccination for homeless | Individuals with ST5 or ST8 IPD were more likely to be homeless (% [OR] 53.0% [4.4] or 43.0% [2.6], respectively). | 0% |  |  | [72] |
| Between 2000-2016 | Outbreak investigation | Calgary, Canada | Hospitals | 1729 patients with confirmed IPD, including 321 HP (18.8%) | Mean±SD: 45.0±27.0 | 78.8% | Culture isolation and serotyping (Gram stain, colony morphology on blood agar, bile solubility, susceptibility to optochin) | The most effective public health intervention for HP to specifically prevent IPD is vaccination (both PPV23 and PCV13 vaccinations) | HP were younger, more often male, smokers, alcohol abusers, illegal drug users, and had a primary diagnosis of pneumonia when compared with non-homeless. | 6.9% |  |  | [73] |
| Between 2000-2014 | Outbreak investigation | Alberta, Canada | Hospitals | 2435 IPD patients, including 184 HP (7.4%) | Mean±SD: 54.2±17.8 | 56.7% | Culture isolation from site such as blood, CSF, pleural fluid, biopsy tissue, joint aspiration, pericardial fluid, or peritoneal fluid | Vaccination and other intervention programmes should have a high benefit in this population. |  | About 15.6% |  |  | [74] |
| Between November 2000 to November 2002 | Outbreak investigation | Alberta, Canada | Hospitals | 129 confirmed BPP patients, including 6 HP (4.7%) | Adults ≥17.0 | About 70% | Culture isolation and serotyping (Gram stain, colony morphology on blood agar, bile solubility, susceptibility to optochin) | HP should be targeted for vaccination. | The attack rate of BPP among HP was 266 per 100,000 person years, 27 times that of the general population. Current smoking, substance abuse, alcohol abuse was predictive of BPP |  |  |  | [75] |
| Between January 2002 and December 2006 | Outbreak investigation | Toronto, Canada | Hospitals | Estimation of 5050 HP in Toronto: 69 (1.4%) cases with confirmed IPD including 27 (0.5%) PP | Mean: 45.0, range: 27.0-74.0 | 89.0% | Culture isolation and serotyping (Gram stain, colony morphology on blood agar, bile solubility, susceptibility to optochin) | Vaccination may be effective in reducing the risk. | HP with IDP were more often smokers, those with HIV infection and liver disease when compared to non-HP | 15.0% |  | Questionnaire: 9.0% | [76] |
| Between 2002-2015 | Outbreak investigation | Anchorage, Alaska, USA | Hospitals | Estimation of 970 HP in Anchorage: 84 cases (8.7%) with confirmed IPD | Mean±SD: 48.0±9.0 | 65.0% | Culture isolation and serotyping | Increasing the availability of pneumococcal vaccine in HP could improve the health of this vulnerable group. | Compared to general population (0.16%), HP were younger, more often alcohol abusers when compared to non-HP. | 0% |  |  | [77] |
| Between April 1988 and March 1989 | Outbreak investigation | Paris, France | Hospitals | 39 HP for acute pneumonia: *Streptococcus pneumoniae* serotype 1, resistant to cotrimoxazole, was isolated in 29 | Mean±SD: 46.0±11.0; range 27.0-74.0 | 100% | Culture isolation and serotyping (Gram stain, colony morphology on blood agar, bile solubility, susceptibility to optochin) | Sheltered residents would benefit from pneumococcal vaccination. |  | 2.6% |  | Questionnaire: 10.0% | [78] |
|  |  |  |  | patients (74.0%). Blood cultures were positive in 24 (61.0%). |  |  |  |  |  |  |  |  |  |
| **B. Diphtheria** |  |  |  |  |  |  |  |  |  |  |  |  |  |
| Between 2016–2017 | Molecular investigation of outbreak | Germany | Hospitals | 76 infected persons, including 25 (32.9%) HP or those reporting drug abuse; strain ST8 (53.9%) | Mean: 45.0 | 83.0% | Culture isolation and genotyping |  |  | 0% |  |  | [79] |
| Between 2004–2012 | Molecular investigation of outbreak | Poland | Hospitals | 18 confirmed diphtheria cases, including 31.0% HP; genotype ST8 (100%) | Range: 16.0–>71.0 |  | Culture, MLST and PFGE |  | Homelessness and alcohol were identified as risk factors. | 0% |  |  | [80] |
| Between 1996–1997 | Outbreak investigation | Switzerland, Germany, France | Hospitals | 17 patients, including 2 HP (11.8%) | Mean: 40.0; range: 30.0–58.0 | 100% | Culture: Isolation from skin or subcutaneous infections |  |  | 0% |  |  | [81] |
| Between 1987–1993 | Outbreak investigation | France | Hospitals | 40 patients with systemic infections, including 13 HP (31.7%) | Mean: 38.0; range: 4.0–87.0 | 75.0% | culture isolation (blood, osteoarticular, perineal, nasopharyngeal, respiratory tract, skin samples) |  | Homelessness and alcohol were identified as risk factors. | 36% |  |  | [82] |
| Between 1972–1982 | Outbreak investigation | Seattle, Washington, USA | Hospitals | 1100 infected persons, including 95.0% HP | Mean: 37.3 (19 HP children + 1081 adults) | About 91.0% | Culture (nasopharyngeal, respiratory tract, skin samples and environmental samples) | Need for vaccination within 10 years | Alcoholic urban HP associated with poor hygiene, crowding, season, contaminated fomites, underlying skin disease, hyperendemic streptococcal pyoderma, and introduction of new strains from exogenous reservoirs. | 0.9% |  |  | [83] |
| **C. Seasonal influenza** |  |  |  |  |  |  |  |  |  |  |  |  |  |
| Between 2017–2018 | Molecular prevalence survey | Marseille, France | Hospital (infectious disease units) | 98 HP hospitalised for different reasons: influenza virus infection (3.0%) | Mean±SD: 43.3 ± 16.8 | 87.6% | qPCR | Need for seasonal flu vaccination |  | 0% |  |  | [15] |
| Between 2015-2017 | Molecular prevalence survey | Marseille, France | Two homeless shelters | 477 HP: Nasopharyngeal influenza carriage (1.7%) | Mean±SD: 43.6 ±16.0; range: 18.0-84.0 | 100% | qPCR | Need for seasonal flu vaccination | Respiratory viral carriage including influenza viruses was positively associated with respiratory symptoms and signs. | 0% | 0.2% were *S. pneumoniae*-influenza virus co-carriage | 15.1% | [2] |
| Between May-November 2009 | Molecular prevalence survey | Tijuana, Mexico | Outreach units and community clinics | 303 persons from marginalised Populations, including HP: H1N1 infection (2.0%) | Mean: 35.0; range: 29.0-43.0 | 62.0% | qPCR | Need for seasonal flu vaccination |  | 0% |  | 8.9% | [84] |
| Year 2005 | Molecular prevalence survey | Marseille, France | Two homeless shelters | 225 HP: Influenza carriage (1.0%) | Mean: 41.0; range: 7.0-76.0 | 94.0% | qPCR | Need for seasonal flu vaccination |  | 0% |  |  | [85] |
| **D. Human papillomavirus** |  |  |  |  |  |  |  |  |  |  |  |  |  |
| Between 2014-2016 | Molecular prevalence survey | Houston, Illinois, USA | Community centres | 130 MSM, including 22.6% HP; 75.0% had at least 1 high-risk HPV type | Range: 18.0-29.0 | 100% | Consensus PCR for HPV genotyping | Black MSM would benefit from increased HPV vaccination efforts |  | 0% |  |  | [86] |
| Between 1995-2002 | Epidemiological survey | New Orleans, Louisiana, USA | Emergency shelter | 4005 HP adolescents: HPV infection (2.0%) among 2723 females | Range: 16.0-21.0 | 32.0% | Pap smear testing |  |  |  |  |  | [46] |

**Abbreviations.** SD, standard deviation; VPD: vaccine-preventable diseases; HP, homeless persons; IPD, Invasive pneumococcal disease; PFGE, Pulsed-field gel electrophoresis; MLST, Multi-locus sequencing typing; MLVA, multiple-locus variable number tandem repeat analysis; CFS, cerebrospinal fluid; OR, Odds-ratio; BPP, Bacteraemic pneumococcal pneumonia; PP, pneumococcal pneumonia; qPCR: real time polymerase chain reaction; HPV, Human papillomavirus; MSM, men who have sex with men.

### Diphtheria (Table 3)

Five diphtheria outbreak investigations were retrieved between 1972 and 2017. Overall, 1251 diphtheria cases were confirmed by culture (with a high prevalence of 12.0%-95.0% cases being homeless persons) [79-83]. Three studies were conducted in adults (mean age: 40-45 years, range 16-71 years) [79-81], and two in both children and adults (mean age: 37-38 years, range 4-87 years) [82, 83]. An early investigation conducted in Seattle, USA between 1972-1982 reported an outbreak of respiratory and cutaneous infections among 1100 patients, including 95% homeless individuals [83]. Molecular studies performed in different types of samples (details not provided) collected in Germany and Poland showed a high 53.9%-100% prevalence of genotype ST8 [79, 80]. One study conducted in Switzerland, Germany and France between 1996-1997 reported 17 cases with cutaneous infection [81]. One study conducted in France between 1987-1993 reported 40 patients with systemic infection [82].

Several risk factors for diphtheria infection were identified, including alcohol abuse [79-81, 83], poor hygiene, crowding conditions, contaminated fomites, underlying skin disease, hyperendemic streptococcal pyoderma, and introduction of new strains from exogenous reservoirs [83]. Death from diphtheria was reported in two studies, with 0.9%-36.0% lethality rates [82, 83]. No data about previous vaccination status was available. One study recommended the need for vaccination boosters in alcoholic homeless persons [83].

### Seasonal influenza (Table 3)

Four PCR-based prevalence surveys conducted among 1103 homeless or persons from marginalised populations between 2005 and 2018 fulfilled the eligibility criteria [2, 15, 84, 85]. Three were conducted among adult sheltered homeless persons in Marseille, France, and one was conducted among marginalised populations, including homeless persons, in Tijuana, Mexico. Three studies were conducted in adults (mean age: 35-44 years, range 18-84 years) [2, 15, 84] and one in a population comprising children and adults (mean age: 41 years, range 7-76 years) [85]. These studies reported a 1.0-3.0% rate of influenza nasopharyngeal carriage among sheltered homeless persons. Using a questionnaire, the previous seasonal influenza vaccination rate among the homeless population was 9.0%-15.0%. The rate of co-infection with *S. pneumoniae* and influenza viruses was 0.2% among homeless individuals in Marseille, France [2]. No influenza deaths were reported. All authors recommended vaccination against influenza for homeless people.

### Human papillomavirus (HPV) infection (Table 3)

Only two American studies concerning HPV infection fulfilled the eligibility criteria. They were conducted in young adults (range 16-29 years) [46, 86]. One molecular prevalence survey was conducted among 130 men having sex with men, of whom 22.6% were homeless and 75% had at least one high-risk HPV type [86]. Another epidemiological study using Pap smear testing in 2960 young female homeless persons revealed a 2.0% rate of HPV infection [46]. Information about vaccination rates was not recorded. No deaths were reported. The vaccination against HPV-16 was recommended for homeless men who have sex with men [86].

### Other VPD

No reports about measles, tetanus, poliomyelitis, pertussis, *H. influenzae* type b infection, meningococcal C infection, mumps, rubella, varicella, chickenpox, typhoid fever, rotavirus infection, cholera, yellow fever, lyssaviruses, rabies, Japanese encephalitis and leptospirosis among homeless people were retrieved.

## 4. Discussion

This review shows that the four most frequent VPD among homeless people are HBV infection, HAV infection, IPD, and diphtheria.

High rates of HBV and HAV infection in homeless persons were observed through serological surveys in low endemicity countries, including the USA, Canada, France and in moderate endemicity countries such as Iran and Brazil. Outbreaks of HAV infection were reported in the US. Currently, three modes of HBV transmission have been recognised: perinatal, sexual and parenteral/percutaneous transmission [87], whilst acute HAV is a highly contagious infection resulting from faeco-oral transmission and has been one of the major aetiologies for foodborne disease [88]. Homeless persons, notably those using injected drugs and having at-risk sexual practices should therefore benefit from vaccination against HBV and HAV infections, together with the provision of injection kits, drug substitutes and condoms.

Outbreaks of IPD were reported in homeless persons in Canada, and several risk factors were identified, including smoking, alcohol abuse, illegal drug use and having several chronic diseases. Given that the serotypes attributable to these outbreaks (1, 4, 5, 8, 12F) are among those included in the 23-valent pneumococcal polysaccharide vaccine, vaccination of homeless people against IPD should be considered, notably when presenting with risk factors.

Finally, because of several diphtheria outbreaks occurring among homeless persons, updating vaccination against this disease should be also considered in this population, as in the general population.

To the best of our knowledge, specific vaccination recommendations for homeless people in national guidelines have been made in only two countries. The 23-valent pneumococcal polysaccharide vaccine (PPSV23) was recommended early in 2008 by the Canadian National Advisory Committee on Immunization (NACI) for all homeless people. In the USA, the adult immunisation schedule [89], updated annually by the Advisory Committee on Immunization Practices (ACIP), has recommended HAV vaccination for all homeless individuals aged > 1 year since October 2018 [3].

In France, the Ministry of Solidarity and Health (Table 4) recommends HAV vaccination for: children aged > 1 year having at least one family member originating from a country with a high prevalence of anti-HAV antibody, for patients with cystic fibrosis or liver disease (notably viral hepatitis and alcohol abuse), for institutionalised infants or young disabled persons, for men having sex with men, for professionals exposed to HAV or involved in collective food preparation and for travellers to endemic areas. For HBV, IPD, and diphtheria, primary vaccination is recommended for all children. The HBV vaccination is notably recommended for institutionalised infants or young disabled persons, for persons in psychiatric institutions, for those with at-risk sexual practices, those using injected drugs or having chronic liver disease or HIV infection, for those with at least one family member with HBV infection, for detained persons, for professionals exposed to HBV (notably health care professionals) and for travellers to endemic areas [90]. PPSV23 together with PCV13 (13-valent pneumococcal conjugate vaccine) is especially recommended for those with several chronic conditions (HIV infection, chronic respiratory disease, heart failure, etc.). No specific recommendations concerning diphtheria vaccination are given for high risk groups other than professionals.

**Table 4.**
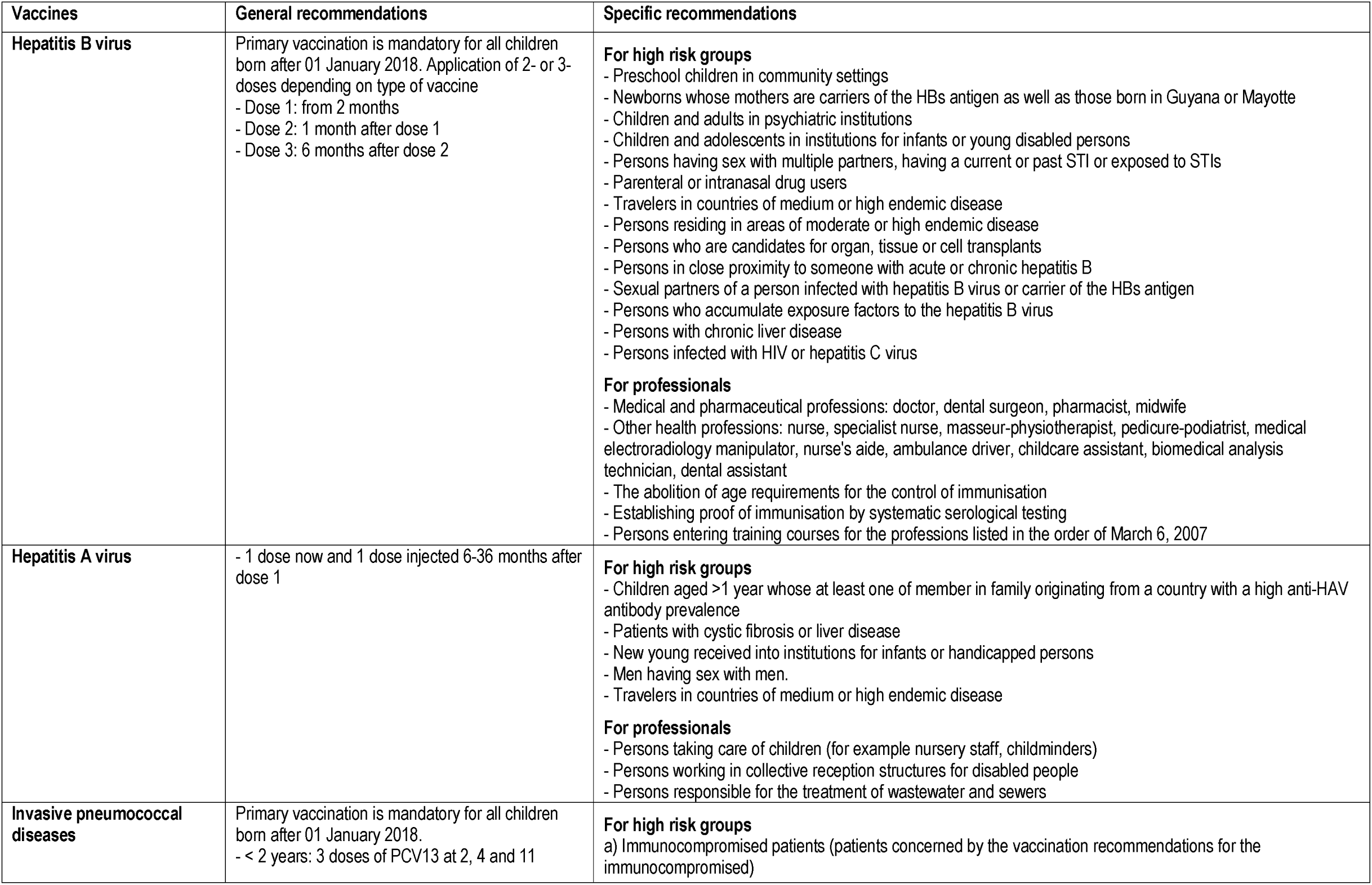
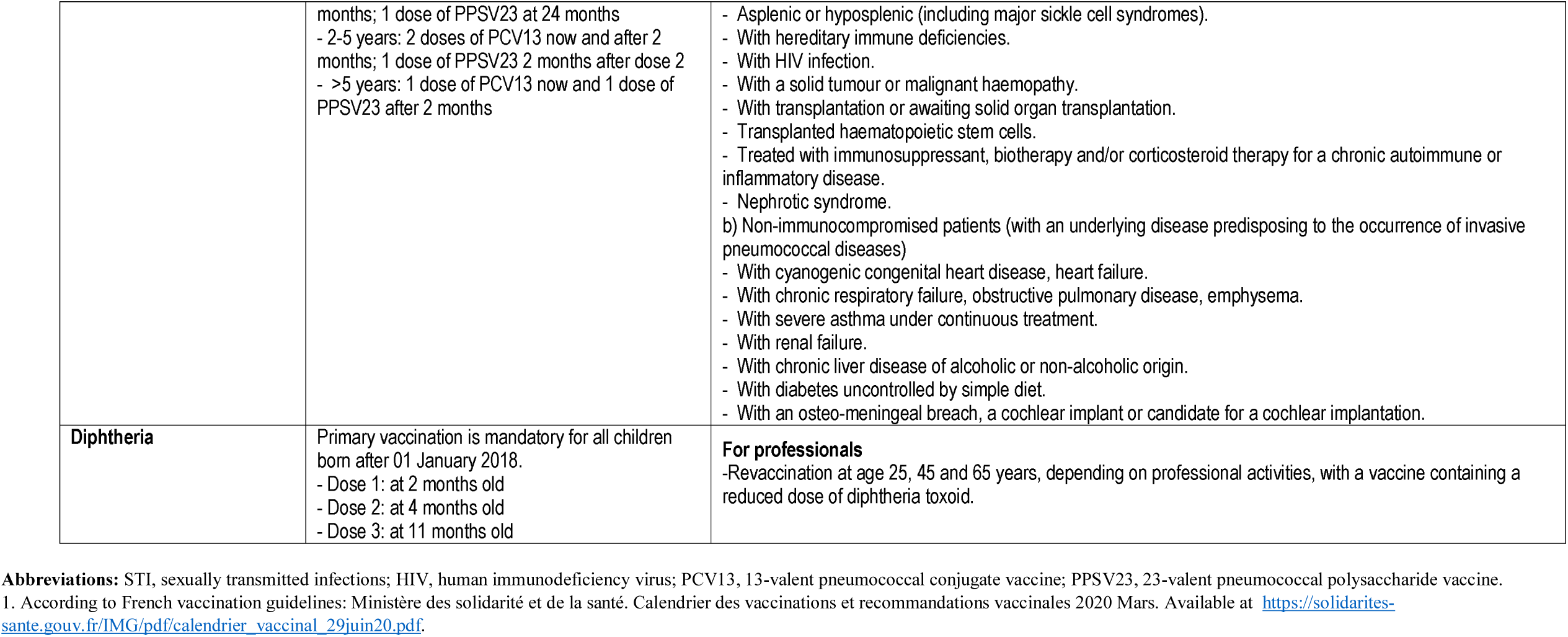
Summary of vaccination against HAV, HBV, pneumococcus and diphtheria for French children and adults^1^

Delivering vaccines to homeless populations has always been a challenge. Combining easy access and education is probably the best approach to vaccination administration programmes in this population. We suggest making vaccines available at homeless shelters or homeless restaurants through medical or paramedical staff. For homeless IDU, the availability of vaccines at syringe-exchange sites could provide more opportunities for them to access vaccination [44]. Additionally, we also suggest applying a rapid serological testing strategy in areas where homeless persons tend to aggregate, including HBsAg detection (for HBV) [91] and IgM/IgG anti-HAV, since a substantial proportion of individuals may already have antibodies against these infections due to past exposure to the viruses or previous vaccination.

Because this review was restricted to articles written in general languages such as English, Spanish and French, the data collected are mainly from the North American and European continents. Limitations include the possible lack of articles from Asian and African regions. Despite these limitations, our review showed that implementation of vaccination coverage against communicable diseases in the homeless is a major public health priority. We strongly recommend updating vaccination against HAV, HBV, pneumococcal infection and diphtheria in homeless persons when entering homeless shelters or whenever they are present in a healthcare setting, rather than as a response measure to prevent outbreaks after the first case has occurred.

## Data Availability

Not applicable

## Author contributions

TD, VT, TL and PG contributed to data analysis, English or French article reading, interpretation and writing. SC contributed to Spanish article reading, interpretation and writing. PG coordinated the work.

