## Supplementary data for "Vaccine-preventable diseases other than tuberculosis, and homelessness: A systematic review of the published literature, 1980 to 2020"

A. Combinations of the following search terms in **Spanish** were used:

#1: ¨Personas en situación de calle¨ OR ¨indigentes¨ OR ¨mendigos¨ OR ¨personas sin hogar¨ OR ¨vagabundos¨

#2: ¨Difteria¨ OR ¨*Corynebacterium diphtheriae*¨ OR ¨tétano¨ OR ¨*Clostridium tetani*¨ OR ¨verrugas genitales¨ OR ¨*Condiloma acuminata*¨ OR¨polio¨ OR ¨poliomyelitis¨ OR ¨poliovirus¨ OR ¨tos ferina¨ OR ¨ toxina Pertussis¨ OR ¨coqueluche¨ OR ¨hepatitis B¨ OR ¨*Haemophilus influenzae*¨ OR ¨meningococo¨ OR ¨*Neisseria meningitidis*¨ OR ¨rubéola¨ OR ¨virus del papiloma humano¨ OR ¨gripa¨ OR ¨gripe¨ OR¨ influenza¨ OR ¨hepatitis A¨OR ¨varicela¨ OR ¨fiebre tifoidea¨ OR ¨fiebre entérica¨ OR ¨Salmonella¨ OR ¨Rotavirus¨ OR ¨Cólera¨ OR ¨*Vibrio cholerae*¨ OR ¨Fiebre amarilla¨ OR ¨Rabia¨ OR ¨*Lyssavirus*¨ OR ¨Encefalitis japonesa¨ OR ¨Leptospira¨ ¨leptospirosis¨

#3: #1 AND #2

B. Combinations of the following search terms in **French** were used:

\# 1: “sans-abri” OR “SDF” OR “sans domicile-fixe ”

\# 2: “diphtérie” OR “*Corynebacterium diphtheriae*” OR “tétanos” OR “*Condyloma accuminata*” OR “polio” OR “poliovirus” OR “poliomyélite” OR “coqueluche” OR “*Bordetella pertussis*” OR “*Hemophilus influenzae* type b“ OR “hépatite B” OR “pneumocoque” OR “*Streptococcus pneumoniae*” OR “ méningocoque” OR “*Neisseria meningitidis*” OR “oreillons “ OR “rougeole” OR “rubéole” OR “papillomavirus humain” OR “influenza” OR “grippe” OR “hépatite A” OR “zona” OR “varicelle” OR “typhoïde” OR “Salmonella” OR “rotavirus” OR “choléra” OR “Vibrio cholerae “ OR “fièvre jaune” OR “Rage” OR “lyssavirus” OR “encéphalite japonaise” OR “leptospirose” OR “Leptospira”

#3: #1 AND #2
